# Metformin Use and Thirty-Day Readmission Among Patients with Bipolar Disorders: A Propensity Score-matching Analysis

**DOI:** 10.1101/2024.07.28.24311148

**Authors:** Ting Zhai

## Abstract

Metformin may affect patients with multi-comorbid bipolar disorder through the integrated stress response. However, the relationship between them remains unclear. This study aimed to explore the relationship between metformin use and early (30-day) readmission in bipolar disorder patients. Data were extracted from the Medical Information Mart for Intensive Care IV database. Adult patients with a documented diagnosis of bipolar disorder were screened. Multivariable logistic regression and propensity score matching were used to investigate potential associations. Data on 1688 patients were included. The crude early (30-day) readmission was significantly higher in patients with metformin use (17/114 vs. 132/1574, *p* = 0.033). In the extended multivariable logistic models, the hazard ratio (HR) of metformin use was consistently significant in all models (HR range 1.87–2.99, *p* < 0.05 for all). After propensity score matching, the early (30-day) readmission remained significantly higher in the metformin use group (Mahalanobis matching, |t|>1.96). Although residual confounding cannot be excluded, metformin use is associated with higher readmission.

## 1. Background

Medical comorbidity is prevalent in bipolar disorder (BD) and may affect its severity and course (Fagiolini et al., 2005), contributing to increased morbidity and mortality (Hajek et al., 2005). Evidence also indicates that a substantial percentage of the cost of BD is due to comorbid, chronic non-communicable diseases that disproportionately affect people with bipolar disorders (e.g., cardiovascular disease) (Kleine-Budde et al., 2014). However, the predominance of recommendations based on opinion beyond the first-line treatment phase reflects the paucity of evidence and the absence of empirical guidance for treating treatment-resistant, tertiary patients with comorbidities (Post et al., 2019).

Considering this reality, the generalisability of much-published research on this issue is problematic. The exclusion of individuals with BD from most pharmacological studies is most commonly encountered in clinical practice (e.g., patients with several comorbidities and suicidality), which will limit the extrapolation of results from these studies to clinical practice (Rosenblat et al., 2019).

New findings (Zhai, 2024) provide further evidence that the integrated stress response (ISR) should be taken seriously because it might improve the treatment response of the management strategies of comorbidities. Some drugs such as metformin, erlotinib, and sunitinib may work through this pathway. However, pathologies such as cognition may have distinct homeostatic set points related to phenotypic fitness. So, either reduced or increased ISR activation can be maladaptive (Costa-Mattioli & Walter, 2020). To further clarify the effects of these drugs, further research is necessary. Due to the problem of data accessibility, we selected metformin as the research object in this study. Moreover, as one of the commonly used drugs to treat diabetes, we don’t know whether long-term use of this drug will have any side effects on patients with bipolar disorder who also often suffer from metabolic diseases. It is hoped that this research will contribute to a deeper understanding of the management of comorbidities in patients with bipolar disorder.

Readmission is common in serious mental illness, leading to increased healthcare costs and adversity for the individual (Schennach et al., 2012). Readmission has been systematically used as a method for measuring the relapse and recurrence of psychiatric illness (Conley et al., 1999) and has often been used as a safe marker of outcomes in psychiatric disorders (Lin et al., 2010). The readmission rate of BD is an important proxy indicator of the frequency of relapse, quality of care, and prognosis, and a major focus of scrutiny for quality improvement for hospitals, health sector administrators, and policymakers (Kristensen et al., 2015). While readmission during a chronic mental illness is not unexpected, it is typically unexpected, particularly within a short timeframe (within 30 days) after discharge, especially if the individual is already engaged with mental health outpatient services (Muhammad et al., 2023).

Therefore, this study aimed to explore the relationship between metformin use and early (30-day) readmission in BD patients, using logistic regression and propensity score matching (PSM).

## 2. Methods

### 2.1. Database introduction

All the data in the current study were extracted from an online international database— Medical Information Mart for Intensive Care IV (MIMIC-IV)—that was published by the Massachusetts Institute of Technology, with approval from the review boards of the Massachusetts Institute of Technology and Beth Israel Deaconess Medical Center (Johnson et al., 2023). All the patients in the database were de-identified for privacy protection, and the need for informed consent was waived. The author obtained access to this database (Record ID: 62627059) and was responsible for data extraction.

### 2.2. Inclusion and exclusion criteria

Adult patients with medical records indicating bipolar disorders. Patients younger than 18 years or whose gender was not recorded were excluded.

### 2.3. Data extraction and outcome definition

Data on the demographic characteristics, laboratory outcomes, and metformin use were extracted from the database. Metformin use was defined as the use of a metformin after admission. Biochemical indicators mainly refer to Surace et al. (2022), including number of white blood cells (WBC) (K/uL), number of red blood cells (m/uL), hemoglobin (HB) (g/dL), number of platelets (K/uL), number of neutrophils (K/uL), number of lymphocytes (K/uL), glucose (mg/dL), blood urea (mg/dL), creatinine (mg/dL), uric acid (mg/dL), aspartate aminotransferase (AST) (IU/L), alanine aminotransferase (ALT) (IU/L), albumin (g/dL), lactate dehydrogenase (LDH) (IU/L), total cholesterol (mg/dL), thyroid-stimulating hormone (TSH) (uIU/mL), and CRP (mg/L).

The primary endpoint was 30-day readmission. Each patient has a unique subject_id, and each admission of the patient will have a unique hadm_id. Based on this index, the admission and discharge time of each record can be used to calculate whether the patient has been readmitted within 30 days.

### 2.4. Propensity score matching

PSM was used to minimize the effect of confounding factors such as hemodynamic indices and disease severity, which may lead to outcome bias. The propensity score was assigned based on the probability that a patient would receive metformin therapy and estimated using a multivariable logistic regression model. The following variables were selected to generate the propensity score: real age, WBC, urea nitrogen, red blood cells, platelet count, HB, and creatinine. Kernel density plots of the p score were used to examine the PSM degree.

### 2.5. Management of missing data

Variables with missing data are common in the MIMIC IV database. For uric acid, TSH, LDH, total cholesterol, CRP, AST, albumin, ALT, absolute neutrophil count, absolute lymphocyte count, and glucose, more than 20% were missing and were removed from this analysis.

### 2.6. Statistical analysis

Continuous variables were presented as mean ± SD or median and interquartile range (IQR) depending on their distribution, as determined by the Shapiro–Wilk test. They were compared using *t-test* for two independent samples or the Wilcoxon rank sum test as appropriate. The Fisher exact test was used to compare categorical data reported as numbers and percentages.

Kaplan–Meier survival curves were generated, and both the Log-rank test and Wilcoxon (Breslow) test were utilized to compare the groups.

A multivariable Exponential regression and Cox proportional hazards model were used to assess the independent association between metformin use and 30-day readmission, after testing the proportional hazards assumption. Results were presented as hazard ratios (HR) with 95% confidence intervals (CI).

PSM was used to minimize the imbalance between groups. All statistical analyses were performed using Stata 16 (Stata Corp., College Station, TX, USA).

## 3. Results

### 3.1. Baseline characteristics

Data on 1688 patients were included. The flow chart of patient selection is presented in Fig. 1. The comparisons of the baseline characteristics are listed in Table 1, after testing, continuous variables all conform to normal distribution. The overall 30-day readmission rate was 8.8%. The difference in Albumin was small between the groups (3.3 ± 0.7 vs. 3.5 ± 0.6, *p* = 0.04). The Red Blood Cells were also relatively similar in the metformin use and no metformin use groups (3.7 ± 0.7 vs. 3.8 ± 0.6, *p* = 0.03), and the Creatinine was similar across the groups despite statistical significance (1.2 ± 1.3 vs. 0.9 ± 0.3, *p* = 0.01). The Urea Nitrogen was significantly higher in patients with metformin use (18.9 ± 14.3 vs. 15.9 ± 7.1, *p* =0.04) while Glucose was lower (123.2 ± 44.5 vs. 168.4 ± 67.8, *p* < 0.01). However, the early (30-day) readmission was significantly higher in patients with metformin use (8.4% vs. 14.9%, *p* = 0.03).

**Figure 1.**
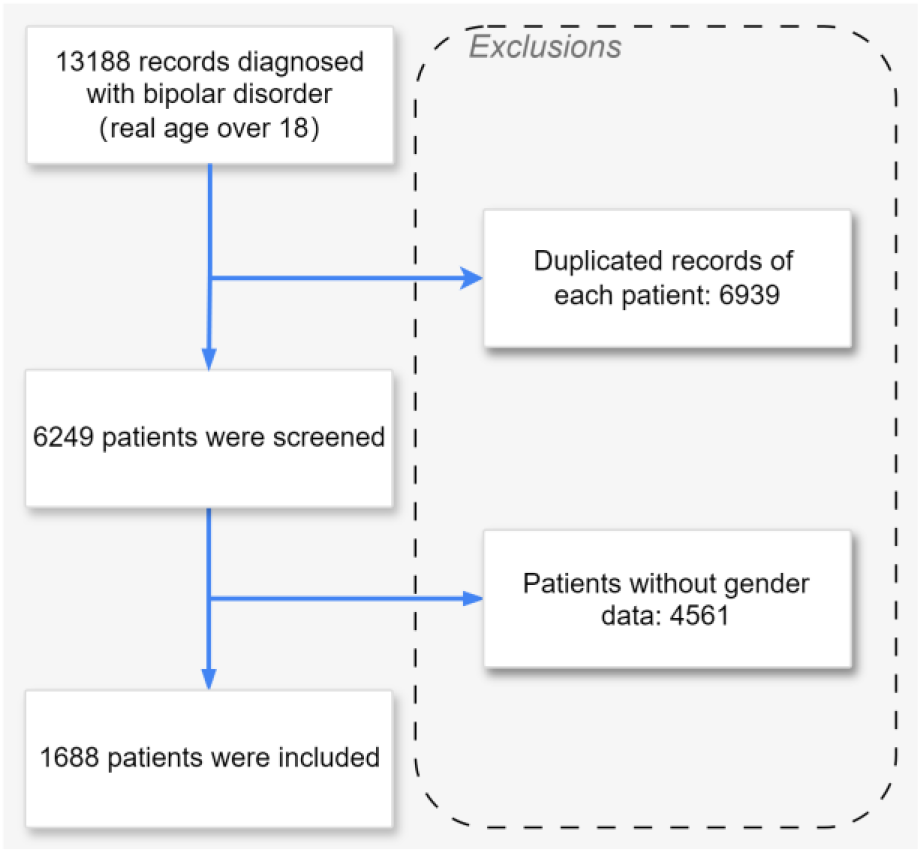
Flow chart of patient selection from the MIMIC IV database.

**Table 1.** Comparisons of the baseline characteristics between patients with and without metformin use.

| Factor | All patients | No metformin use | Metformin use | p-value | Test |
| --- | --- | --- | --- | --- | --- |
| N | 1688 | 1574 | 114 |  |  |
| Male | 843 (49.9%) | 788 (50.1%) | 55 (48.2%) | 0.77 | Fisher's exact |
| Real_age*, mean (SD) | 52.5 (16.0)<br>(n=1688) | 52.1 (16.1)<br>(n=1574) | 57.4 (13.6)<br>(n=114) | <0.01 | Two sample t test |
| Race |  |  |  | <0.01 | Fisher's exact |
| PATIENT DECLINED TO ANSWER /UNABLE TO OBTAIN/UNKNOWN | 271 (16.1%) | 262 (16.6%) | 9 (7.9%) |  |  |
| HISPANIC/LATINO | 59 (3.5%) | 54 (3.4%) | 5 (4.4%) |  |  |
| BLACK | 178 (10.5%) | 157 (10.0%) | 21 (18.4%) |  |  |
| WHITE | 1126 (66.7%) | 1053 (66.9%) | 73 (64.0%) |  |  |
| ASIAN | 7 (0.4%) | 6 (0.4%) | 1 (0.9%) |  |  |
| NATIVE HAWAIIAN OR OTHER PACIFIC ISLANDER | 3 (0.2%) | 2 (0.1%) | 1 (0.9%) |  |  |
| AMERICAN INDIAN/ALASKA NATIVE | 2 (0.1%) | 1 (0.1%) | 1 (0.9%) |  |  |
| MULTIPLE RACE/ETHNICITY | 3 (0.2%) | 3 (0.2%) | 0 (0.0%) |  |  |
| OTHER | 39 (2.3%) | 36 (2.3%) | 3 (2.6%) |  |  |
| Biochemical indices |  |  |  |  |  |
| Thyroid Stimulating Hormone, mean (SD) | 3.5 (6.4) (n=372) | 3.6 (6.7) (n=339) | 2.1 (1.6) (n=33) | 0.21 |  |
| Lactate Dehydrogenase (LD), mean (SD) | 424.2 (790.1) (n=651) | 435.9 (810.9) (n=615) | 224.2 (110.3) (n=36) | 0.12 |  |
| Cholesterol, Total, mean (SD) | 155.7 (55.1) (n=127) | 154.9 (56.6) (n=113) | 161.8 (42.4) (n=14) | 0.66 |  |
| C-Reactive Protein, mean (SD) | 64.4 (71.9) (n=93) | 64.3 (72.1) (n=85) | 65.3 (74.8) (n=8) | 0.97 |  |
| Asparate Aminotransferase (AST), mean (SD) | 166.1 (735.0) (n=961) | 174.1 (757.1) (n=904) | 38.9 (32.6) (n=57) | 0.18 |  |
| Albumin, mean (SD) | 3.3 (0.7) (n=724) | 3.3 (0.7) (n=682) | 3.5 (0.6) (n=42) | 0.04 |  |
| Alanine Aminotransferase (ALT), mean (SD) | 154.0 (597.0) (n=958) | 161.4 (614.7) (n=901) | 37.1 (39.8) (n=57) | 0.13 | Two sample t test |
| Absolute Neutrophil Count, mean (SD) | 8.3 (5.9) (n=378) | 8.3 (6.0) (n=349) | 7.3 (4.5) (n=29) | 0.38 |  |
| Absolute Lymphocyte Count, mean (SD) | 41.9 (261.3) (n=385) | 42.7 (268.0) (n=356) | 31.9 (160.2) (n=29) | 0.83 |  |
| Glucose, mean (SD) | 126.2 (47.7) (n=1477) | 123.2 (44.5) (n=1379) | 168.4 (67.8) (n=98) | <0.01 |  |
| White Blood Cells, mean (SD) | 9.3 (6.1) (n=1469) | 9.3 (6.3) (n=1373) | 9.2 (3.5) (n=96) | 0.85 |  |
| Urea Nitrogen, mean (SD) | 18.7 (13.9) (n=1479) | 18.9 (14.3) (n=1381) | 15.9 (7.1) (n=98) | 0.04 |  |
| Red Blood Cells, mean (SD) | 3.7 (0.7) (n=1469) | 3.7 (0.7) (n=1373) | 3.8 (0.6) (n=96) | 0.03 |  |
| Platelet Count, mean (SD) | 228.0 (104.9)<br>(n=1469) | 227.3 (105.9)<br>(n=1373) | 237.1 (89.2)<br>(n=96) | 0.38 |  |
| Hemoglobin, mean (SD) | 11.0 (2.0)<br>(n=1470) | 11.0 (2.0)<br>(n=1374) | 11.2 (1.8)<br>(n=96) | 0.48 |  |
| Creatinine, mean (SD) | 1.2 (1.2)<br>(n=1482) | 1.2 (1.3)<br>(n=1384) | 0.9 (0.3)<br>(n=98) | 0.01 |  |
| Readmission_30days | 149 (8.8%) | 132 (8.4%) | 17 (14.9%) | 0.03 | Fisher's exact |

\* Real age = anchor\_age + admittance - anchor\_year
Note: Uric Acid and CRP were not recorded in the cases that met the inclusion criteria.

### 3.2. Association between metformin use and 30-day readmission

#### 3.2.1 Survival analysis

With the Log-rank test for equality of survivor functions (*p* = 0.0109) and Wilcoxon (Breslow) test for equality of survivor functions (*p* = 0.0108), we reject the null hypothesis (there is no significant difference between the two drug groups). Figure A1 demonstrated that the metformin group did have a more severe time-to-readmission profile.

#### 3.2.2 Exponential regression

First, we perform the simplest single-parameter exponential regression. The results showed that the risk ratio of metformin use was about 2.990, which means that metformin users have a 199% higher risk of 30-day readmission than non-users. Since the risk function of exponential regression is a constant, the assumption is too strong, so we relax this assumption and perform Weibull regression.

The P value corresponding to the original hypothesis “*H*_0_: lnp = 0” is < 0.0001, so the exponential regression is strongly rejected, and it is believed that Weibull regression should be used. Furthermore, 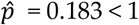, so the risk function decreases over time, and there is a negative duration dependence.

Next, we conduct another generalization of exponential regression, namely Gompertz regression. The P value corresponding to the original hypothesis “H0: lnp = 0” is < 0.0001, so the exponential regression is strongly rejected, and it is believed that Gompertz regression should be used. As for choosing between Weibull regression and Gompertz regression, it can be done according to the AIC criterion. Since the number of explanatory variables and parameters of these two regressions are equal, the AIC criterion is equivalent to maximizing the log-likelihood function. Since the log-likelihood value of Weibull regression is -786 and the log-likelihood value of Gompertz regression is -610, Gompertz regression should be selected.

Other candidate parametric regressions include lognormal regression and loglogistic regression belonging to the accelerated failure time model (AFT). Lognormal regression is performed first. The coefficients of the lognormal regression have the opposite sign to those of the proportional hazard model because they represent the semi-elasticity of the variable to the mean life span, while the coefficients in the proportional hazard model represent the semi-elasticity to the hazard rate. The higher the hazard rate, the shorter the mean life span; therefore, the two have opposite effects. The log-likelihood value of the lognormal regression is -775, slightly inferior to that of the Gompertz regression.

Next, we perform logistic regression. The log-likelihood value of logistic regression is -784, which is also lower than that of Gompertz regression.

#### 3.2.3 Cox regression

Since we are still unsure about the specific distribution of parametric regression, we perform semi-parametric Cox regression below (Model 1, Model 2, and Model 3). From Table A1, we can see that overall, the assumption of proportional hazards is acceptable. However, the variable Hemoglobin may violate the proportional hazards assumption, with a P value of 0.0339.

We test the proportional hazards model by introducing time-varying explanatory variables (TVC). Specifically, when the interaction term between the variable Hemoglobin and time t is introduced, Cox regression is performed, and then the significance of this interaction term is tested. The p-value of the interaction term Hemoglobin×t is 0.217, so the interaction term can be considered to be insignificant at the 5% level, supporting the proportional hazard assumption.

In the multivariable logistic models (Table 2), we observed that the hazard ratio (HR) of metformin use was consistently significant in all models (HR range 1.87–2.99, p < 0.05 for all). A positive correlation was found between metformin use and readmission.

**Table 2.**
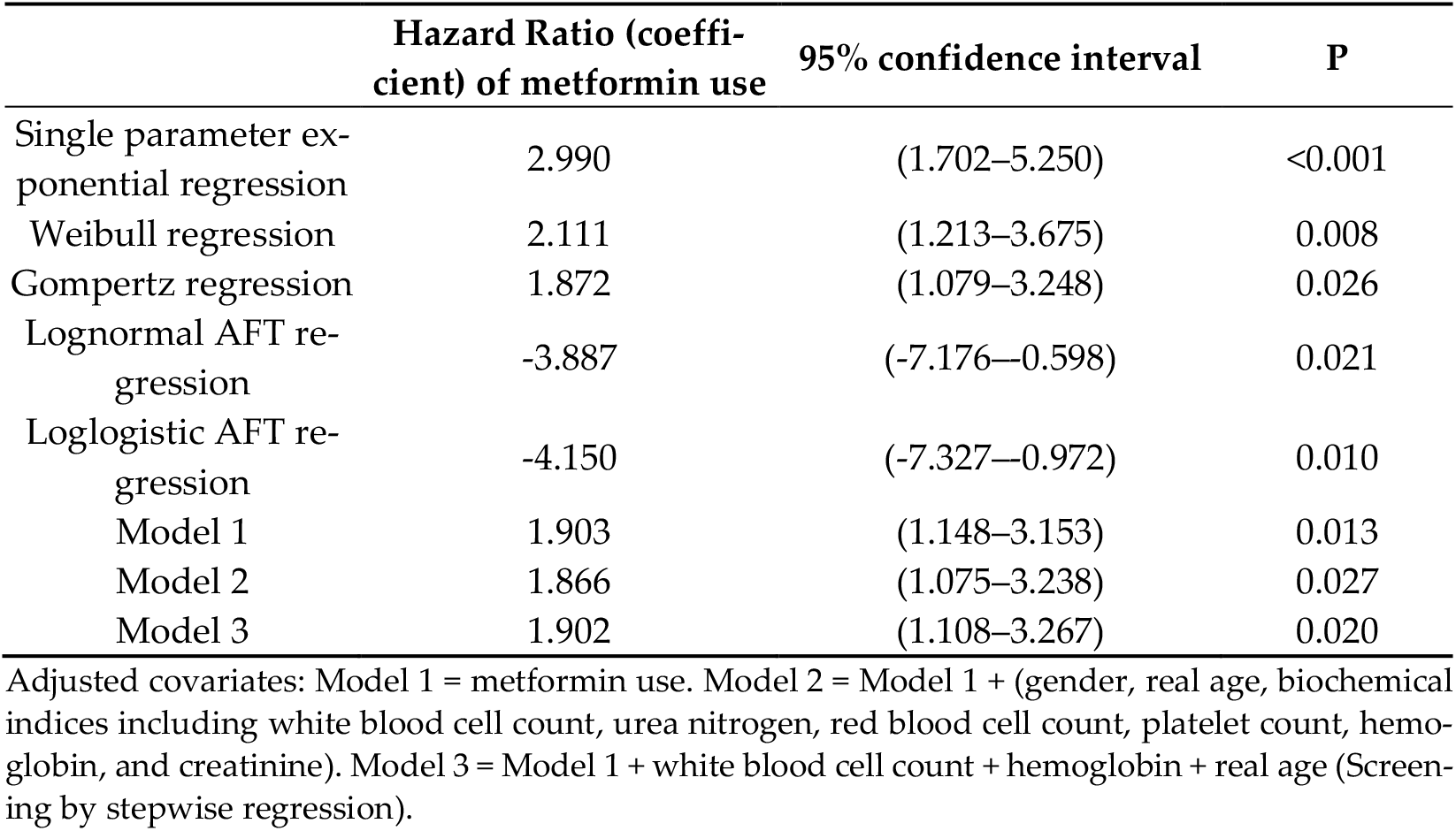
Association between metformin use and 30-day readmission.

### 3.3. Outcomes after propensity score matching

The following is a one-to-one matching. Since the sample size is not very large, a matching with replacement is performed, and parallel.

The estimated value of ATT is 0.053, and the corresponding t value is 1.03 which is less than the critical value of 1.96, so it is insignificant. Among the total 1454 observations, 151 in the control group (Untreated) are not in the common value range (offsupport), and 0 in the treatment group (Treated) are not in the common value range (offsupport). The remaining 1303 observations are all in the common value range (onsupport).

Table A2 shows that the standardized deviations of most variables after matching are less than 10%, except for the variable red blood cells and HB, which has a deviation of 14.8% and 14.7%, which seems to be acceptable; and the results of most t-tests do not reject the original hypothesis that there is no systematic difference between the treatment group and the control group. Comparing the results with the pre-matching results, the standardized deviations of most variables were substantially reduced, but the deviations of the variables white blood cells and HB increased instead. From Figure A2, we can intuitively see that the standardized deviations of most variables are reduced after matching and most observations are within the common range (on support), so only a small number of samples will be lost when performing propensity score matching.

In summary, As can be seen from Table 3, the various propensity score matching results show that the average treatment effect of metformin use is positive and statistically significant (Most are significant at the 10% level, |t|>1.64).

**Table 3.** ATT of propensity score matching results.

|  | Difference | S.E. | T-stat |
| --- | --- | --- | --- |
| 1:1 matching | 0.053 | 0.052 | 1.03 |
| 1:10 matching | 0.077 | 0.040 | 1.91 |
| 1:10 matching in the caliper* | 0.076 | 0.040 | 1.88 |
| Radius (caliper) matching | 0.064 | 0.040 | 1.61 |
| Kernel matching (using default kernel function and bandwidth) | 0.062 | 0.039 | 1.58 |
| Local linear regression matching (using default kernel function and bandwidth) | 0.068 | 0.040 | 1.70 |
| Mahalanobis matching | 0.088 | 0.040 | 2.18 |

\* $0.25\hat{\sigma}_{pscore} = 0.01038327$ . To be conservative, the caliper range is set to 0.01, which means that one-to-10 matching is performed for observations with a 1% difference in propensity scores.

## 4. Discussion

The present study demonstrates that the use of metformin in BD patients is associated with significantly increased early (30-day) readmission. This result was robust in the PSM analysis after adjustment for covariates.

From the perspective of BD itself, this finding is contrary to previous studies which have suggested that metformin may have a protective effect on cognitive and neurodegenerative disorders through the PERK (Chen et al., 2018; Kumar et al., 2023). Given that myelinating cells from either the central or the peripheral nervous system synthesize a large number of myelin proteins and lipids, they typically accumulate misfolded or unfolded proteins and activate both the unfolded protein response (UPR) and the ISR. Many studies have postulated a convergence between bipolar disorder and abnormal myelination (Chambers & Perrone-Bizzozero, 2004; Marlinge et al., 2014; Tkachev et al., 2003; Walterfang et al., 2009; Xu et al., 2022). In myelination disorders, the role of the ISR in pathology is complex. In some conditions, activation of the ISR is protective: Whereas GCN2 protects oligodendrocytes and white matter during branch amino acid deficiency (She et al., 2013), PERK protects from demyelination and axonal degeneration in a mouse model of experimental autoimmune encephalomyelitis (Lin et al., 2007). Correspondingly, in mouse models of Charcot-Marie-Tooth disease that exhibit increased eIF2-P concentrations, germline ablation (D’Antonio et al., 2013; Pennuto et al., 2008) or pharmacological inhibition of GADD34 with sephin1 improved motor function and reduced demyelination (Das et al., 2015). Counterintuitively, in the same model, removing one copy of PERK in all or just in Schwann cells partially reversed the pathology (Musner et al., 2016; Sidoli et al., 2016). Thus, inhibition of the ISR could have disease-protective or disease-causing effects. However, the molecular mechanisms underlying these two opposing outcomes remain unclear. However, for this article, this observation may support the hypothesis that metformin blocks the inhibitory effect of extracellular ATP on Schwann cell proliferation and differentiation by antagonizing lysosomal exocytosis (Jung et al., 2013; Shin et al., 2012). Schwann cells highly express P0 and P0S63del mRNA and protein at P28 (Wrabetz et al., 2006) and persistently activate PERK, which has detrimental effects on nerves (Musner et al., 2016).

What’s more, from the perspective of metabolic disorders, metformin is a well-known drug for the treatment of diabetes and is widely used in clinical practice. However, permanent activation of the ISR, as determined by increased eIF2-P, is also not tolerated by pancreatic β-cells. Patients carrying a specific mutation in eIF2(γ), which results in a decrease in ternary complex (TC) and translation fidelity, exhibit hyperglycemia and diabetes (Moortgat et al., 2016; Stanik et al., 2018). Accordingly, reduced TC owing to a loss-of-function mutation in CReP has been associated with diabetes (Abdulkarim et al., 2015). CReP-deficient β-cells are more susceptible to apoptosis: It seems that persistent activation of the ISR and ATF4 translation induces the transcription of the proapoptotic transcription factor CHOP, which promotes apoptosis (Zinszner et al., 1998). Thus, in β-cells, precise ISR-mediated translation regulation is important for adequate ER stress management. The observation that both enhancement and inhibition of the ISR lead to pathology indicates that normal β-cell function requires eIF2-P abundance to be maintained in a narrow range and/or that the ISR needs to switch on and off dynamically. Compound-based strategies that compensate for deviating eIF2-P abundance by adjusting the extent of ISR activation or inhibition will be required to improve the pathology associated with these disorders. Thus, depending on the disease, either ISR inhibition or ISR activation may be beneficial therapeutically. However, also due to metformin inhibits mitochondrial ATP production, and it is essential for β-cell insulin secretion (Lamontagne et al., 2017). Plus, prior studies have noted that metformin is proposed to potentially affect β-cells, especially at high circulating levels (Valle et al., 2022).

This study has several limitations. First, there are many confounders for metformin use in clinical practice, such as the disease severity and clinician preference. In the current study, we included as many confounders as possible to minimize potential bias. However, due to the retrospective nature, some information was unavailable in this database. Rigorously designed randomized controlled studies may be the only solution for the imbalance between these two groups. Second, we did not consider pre-admission mental disorder severity, medication compliance, and the dose/duration of individual metformin used in the hospital, and further research is necessary to confirm any association. Third, our study only included patients from a single center, which limits its generalizability to other hospitals with varying practices or resources. Finally, the causal relationship between metformin use and early (30-day) readmission could not be confirmed. While the use of a propensity score can further support our speculation, it still cannot overcome the primary limitation associated with the observational nature of the study. Further prospective studies are needed to verify our hypothesis.

## 5. Conclusions

The metformin use was associated with an increased risk of readmission and mortality. This emphasizes that drug treatment for BD patients should be combined with the management of comorbidities and taking preventative measures to manage the potential side effects of combined medication to reduce readmission. Future larger randomized clinical trials are required to confirm and validate this association.

## Author Contributions

All work was done by the only author Zhai Ting.

## Funding

This research received no external funding

## Data Availability Statement

The datasets presented in the current study are available in the MIMIC-IV database (https://physionet.org/content/mimiciv/2.2/).

## Acknowledgments

The author thanks Prof. Deng Huihua for his help in critically discussing and reviewing the manuscript.

## Conflicts of Interest

The authors declare no conflicts of interest.

## Appendix A

**Figure A1.**
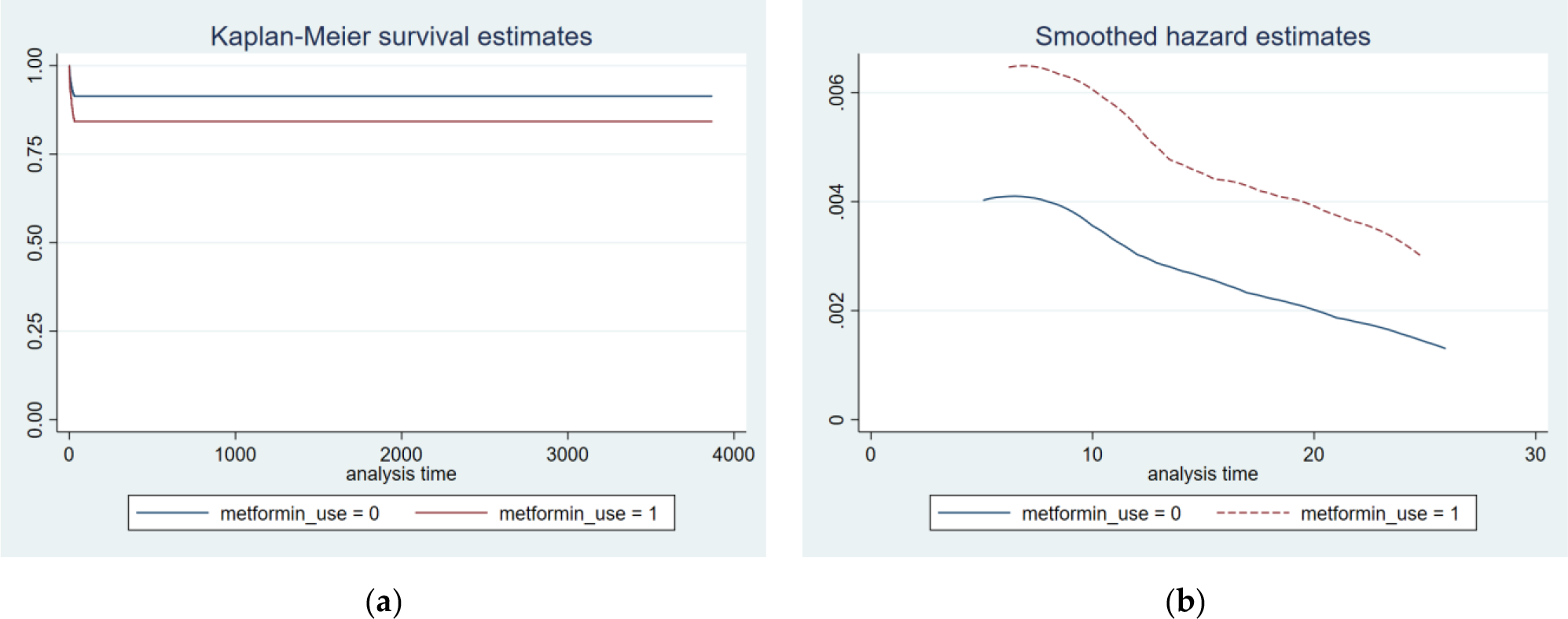
(**a**) Kaplan–Meier curves for the metformin use; (**b**) Risk function according to metformin use grouping.

**Figure A2.**
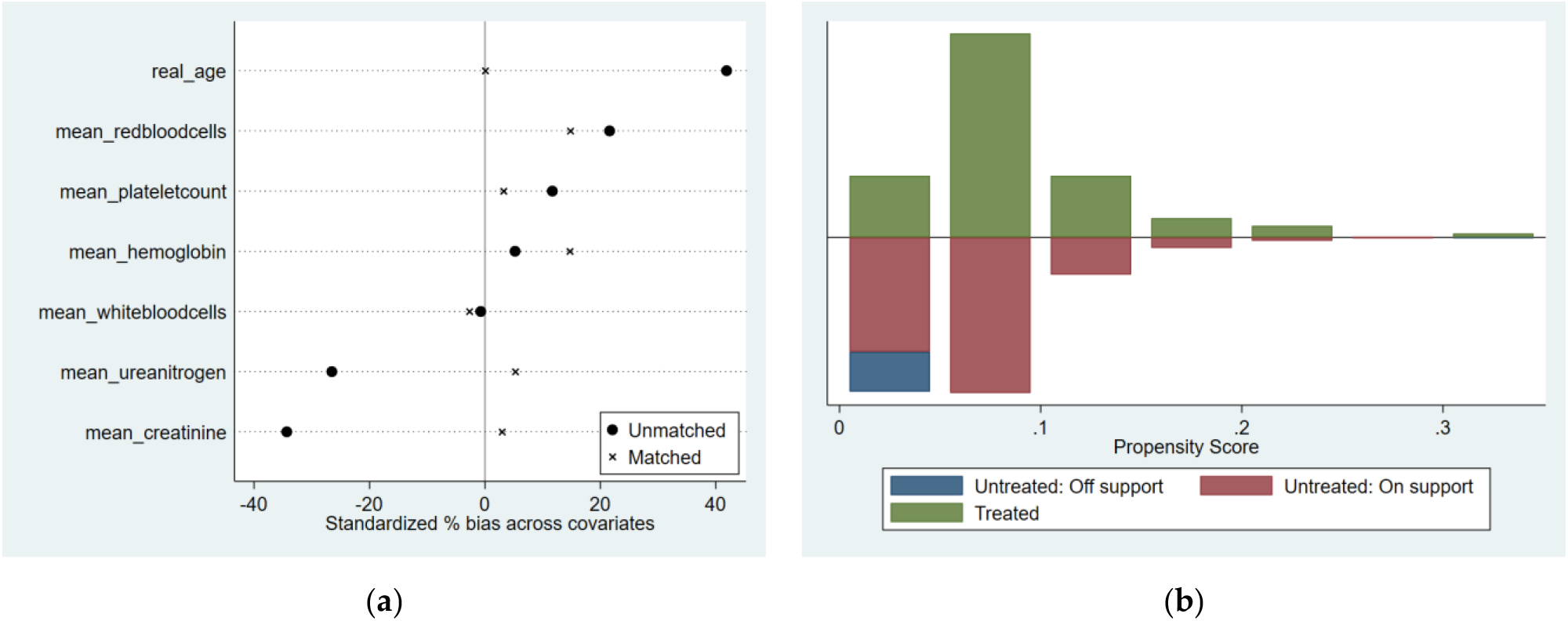
(**a**) Graphical representation of standardized deviations for each variable; (**b**) Common range of propensity scores.

**Figure A3.**
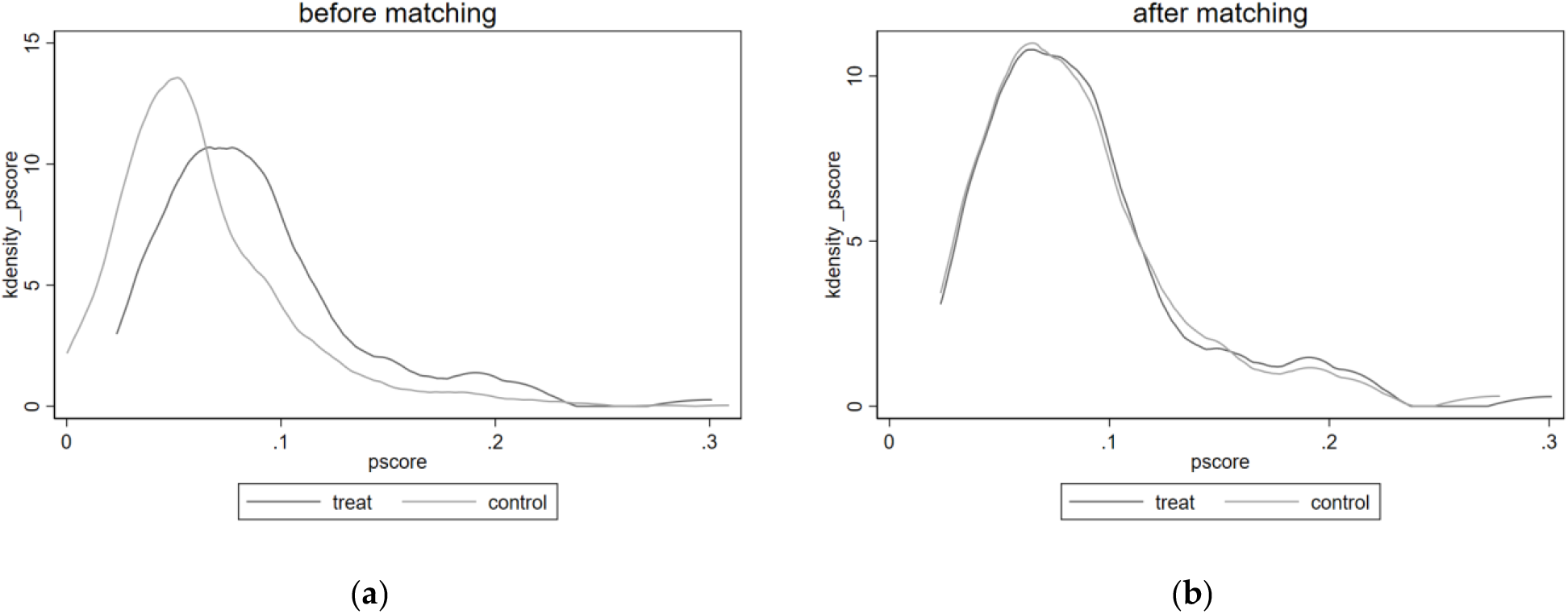
Kernel density plots of the propensity score before and after propensity score matching.

**Table A1.** P value of test of proportional-hazards assumption.

|  | Model 1 | Model 2 | Model 3 |
| --- | --- | --- | --- |
| Metformin use | 0.9605 | 0.6039 | 0.8380 |
| male | NA | 0.5441 | NA |
| WBC | NA | 0.7483 | 0.3883 |
| Urea Nitrogen | NA | 0.8539 | NA |
| RBC | NA | 0.0548 | NA |
| Platelet Count | NA | 0.5019 | NA |
| Hemoglobin | NA | 0.0339 | 0.2842 |
| Creatinine | NA | 0.8978 | NA |
| Real age | NA | 0.3169 | 0.4316 |
| NA not applicable |  |  |  |

**Table A2.** Equilibrium metrics after matching.

| Variable | Unmatched<br>Matched | Mean |  | %bias | %reduct<br>\|bias\| | t-test |  | V(T)/<br>V(C) |
| --- | --- | --- | --- | --- | --- | --- | --- | --- |
|  |  | Treated | Control |  |  | t | p>\|t\| |  |
| White blood cells | U | 9.2287 | 9.2652 | -0.7 |  | -0.06 | 0.955 | 0.32* |
|  | M | 9.2287 | 9.3641 | -2.7 | -271 | -0.25 | 0.802 | 0.82 |
| Urea nitrogen | U | 15.878 | 18.875 | -26.5 |  | -2.02 | 0.044 | 0.25* |
|  | M | 15.878 | 15.282 | 5.3 | 80.1 | 0.58 | 0.564 | 1.08 |
| Red blood cells | U | 3.8072 | 3.6717 | 21.6 |  | 1.93 | 0.054 | 0.79 |
|  | M | 3.8072 | 3.7143 | 14.8 | 31.4 | 1.07 | 0.285 | 0.96 |
| Platelet count | U | 238.59 | 227.16 | 11.7 |  | 1.02 | 0.30-8 | 0.7 |
|  | M | 238.59 | 235.43 | 3.2 | 72.3 | 0.20 | 0.843 | 0.49* |
| hemoglobin | U | 11.105 | 11.006 | 5.2 |  | 0.47 | 0.636 | 0.85 |
|  | M | 11.105 | 10.826 | 14.7 | -181.6 | 1.05 | 0.297 | 0.97 |
| creatinine | U | 0.88351 | 1.2056 | -34.3 |  | -2.41 | 0.016 | 0.05* |
|  | M | 0.88351 | 0.85565 | 3 | 91.3 | 0.64 | 0.521 | 0.99 |
| Real age | U | 59.372 | 53.144 | 41.9 |  | 3.66 | <0.001 | 0.71 |
|  | M | 59.372 | 59.362 | 0.1 | 99.8 | 0.01 | 0.996 | 1.18 |

## Notes

### Competing Interest Statement

The authors have declared no competing interest.

### Funding Statement

This study did not receive any funding

### Summary of Updates

Supplemental files updated.Some grammatical errors were fixed。

